# Exploring associations between estrogen and gene candidates identified by coronary artery disease genome-wide association studies

**DOI:** 10.1101/2024.08.29.24312812

**Authors:** Ava P. Aminbakhsh, Emilie T. Théberge, Elizabeth Burden, Cindy Kalenga Adejumo, Anna Lehman, Tara L. Sedlak

## Abstract

**Introduction:** Coronary artery disease (CAD) is the leading cause of death around the world, with well-described epidemiological sex and gender differences in prevalence, pathophysiology and management outcomes. It has been hypothesized that sex steroids, like estrogen, may contribute to these sex differences. There is a relatively large genetic component to developing CAD, with heritability estimates ranging between 40-60%. In the last two decades, the computational methods, capabilities and scalability of genome-wide association studies (GWAS) have contributed substantially to advancing the understanding of which genetic candidates contribute to CAD. The aim of this study was to determine if genes discovered in CAD GWASs are affected by estrogen by means of direct modulation or indirect down-stream targets.

**Methods:** A scoping review of the literature was conducted using MEDLINE and EMBASE through to April 24, 2024, for studies synonymous to an atherosclerotic coronary artery disease phenotype, and a genome-wide association study (GWAS) design. Analysis was limited to candidate genes with corresponding single nucleotide polymorphisms (SNPs) surpassing genome-wide significance and had been mapped to genes by study authors. The number of studies that conducted sex-stratified analyses with significant genes were quantified. A literature search of the final gene lists was done to examine any evidence suggesting estrogen may modulate the genes and/or gene products.

**Results:** There were 60 eligible CAD GWAS studies meeting inclusion criteria for data extraction. Of these 60, only 36 had genome-wide significant SNPs reported, and only 3 of these had significant SNPs from sex-stratified analyses mapped to genes. From these 36 studies, a total of 61 genes were curated, of which 26 genes (43%) were found to have modulation by estrogen. All 26 were discovered in studies that adjusted for sex. 12/26 genes were also discovered in studies that conducted sex-stratified analyses. 12/26 genes were classified as having a role in lipid synthesis, metabolism and/or lipoprotein mechanisms, while 11/26 were classified as having a role in vascular integrity, and 3/26 were classified as having a role in thrombosis.

**Discussion:** This study provides further evidence of the relationship between estrogen, genetic risk and the development of CAD. More sex-stratified research will need to be conducted to further characterize estrogen’s relation to sex differences in the pathology and progression of CAD.

## Introduction

Coronary artery disease (CAD) is the leading cause of death around the world (1), with well-described epidemiological sex and gender differences in prevalence, pathophysiology, and management outcomes (2) (Box 1). CAD is attributed to atherosclerosis, where lesions (atheromas or atherosclerotic plaques) form in the luminal intima of coronary arteries. These lesions develop over time due to a complex interplay of risk factors within a state of chronic inflammation (3). Sex differences have been studied in the development and progression of atherosclerosis but are not completely understood (4). For instance, cisgender women have been found to have smaller coronary arteries and are more likely to experience plaque erosion, whereas men are more prone to plaque rupture (5).

Sex differences in the presentation of CAD include the higher prevalence of traditional cardiovascular risk factors and a later average onset in women, approximately 10 years later than in men (6). Higher sex-specific risk in developing CAD has been observed in women who have diabetes, a history of smoking, depression and/or anxiety. Unique risk factors of women include sex-specific conditions such as premature menopause, gestational hypertension or preeclampsia, and polycystic ovarian syndrome (PCOS) (7–9). In pre- and peri-menopausal populations of women, the risk of CAD is perceived to be lower due to the protective role of estrogens (10).

Cardiovascular health is modulated in part by the steroid sex hormones: androgens, estrogens and progestogens, among which the most abundant subtypes are testosterone, 17β -estradiol and progesterone, respectively (6,11). These hormones bind to receptors expressed on the surface of most cell types in the cardiovascular system in both sexes, exerting physiological effects through regulation of genomic expression (“slow response”, spanning hours to days), or acting on extranuclear components in the cell (“fast non-genomic” response, spanning seconds to minutes) (12,13). Estrogen has been shown to exert both direct and indirect regulatory effects on thousands of genes (14). Protective physiological effects include the promotion of vasodilation, anti-inflammatory cascades, and improvement of lipid profiles such as through decreased low-density lipoprotein (LDL) oxidation and binding (15–18). Theoretically, deleterious genetic variations in genes regulated by estrogen and with implications in cardiovascular health could increase the predisposition for CAD.

There are sex differences in physiologically normal ranges of androgen to estrogen ratios, which further differ by age (i.e., puberty, menopause, andropause) (19,20). In women, premenopausal estradiol ranges from 30 to 400 pg/mL while postmenopausal estradiol ranges from 0 to 20 pg/mL (21,22), while androgen levels mostly constant but gradually decrease over the course of a lifetime, and not to the rate or degree of estrogen decline (23). As such, the ratio of androgen to estrogens differs greatly between pre and post-menopausal women (19,20). When levels go outside normal ranges, variable effects are observed on cardiovascular health. Women with PCOS, a condition characterized by elevated androgen levels that disrupt the estrogen-to-testosterone ratios, have been shown to experience accelerated atherosclerosis compared to women without PCOS (24,25). Further, a number of candidate gene studies have identified associations with increased risk of CAD or myocardial infarction (MI) risk in individuals with deleterious variants in the estrogen receptor 1 (*ESR1* or *ERα,*) or 2 genes (*ESR2* or *ERβ*) causing dysfunctional or null activity (26–29).

There is a relatively large genetic component to developing CAD, with heritability estimates ranging between 40-60% (30,31). Most of the heritability of CAD is polygenic, owing to the individually small but cumulatively large contribution of hundreds to thousands of genes considered in CAD risk (32). In the last two decades, the computational methods, capabilities and scalability of genome-wide association studies (GWAS) have contributed substantially to advancing the understanding of which genetic candidates contribute to CAD. Following the first CAD GWAS published in 2007 (33), novel or overlapping candidate genes have emerged through replication between studies, with differences between studies reflecting parameters such as sample size, genetic ancestry, male/female representation, and CAD definitions. Historically, GWASs were predominantly in populations of European genetic ancestry; with time, more diverse and multi-ethnic cohorts have emerged to broaden the generalizability and unique candidates from GWAS findings across different population groups. Furthermore, in addition to the small effects of many common genetic variants towards CAD risk, there are also genes that can contribute large independent risk towards CAD if an individual carries deleterious variants, such as in *LDLR, APOB, PCSK9*, and *LPA* (34,35).

Investigating sex as a biological variable in GWAS studies is essential for accurately identifying sex-specific genetic associations and understanding their unique contributions to disease susceptibility through affected biological pathways. Studies that do not segregate by sex assume that the contributing genes to biological pathways of disease are shared between the sexes; this reduces trait specificity and inserts a level of bias into the results (36). The rationale may be from a fear of potential lowered statistical power when sample sizes are sex-stratified; however, this operates under the assumption that there are no sex differences. If there are indeed sex differences, sex-stratified analyses could in fact increase the power to detect these differences.

To our knowledge, there has not been a review conducted to date that quantifies CAD GWAS-identified gene candidates from both a sex-stratified lens and in assessment of associated modulation by estrogens. Therefore, the aims are twofold: first, we aim to quantify the number of CAD GWAS studies published to date that conducted any sex-stratified analyses. Second, we aim to identify gene candidates listed in both sex-stratified and non-stratified CAD GWAS publications that have functional evidence for direct or indirect regulation by estrogens.

### Box 1

Throughout this review, the terms sex and gender are used interchangeably due to inconsistent terminology used throughout the studies. These terms are distinguished by the Canadian Institutes for Health Research Panel on Sex and Gender (37) as follows: “Sex refers to a set of biological attributes in humans and animals. It is primarily associated with physical and physiological features including chromosomes, gene expression, hormone levels and function, and reproductive/sexual anatomy. Sex is usually categorized as female or male but there is variation in the biological attributes that comprise sex and how those attributes are expressed. Gender refers to the socially constructed roles, behaviours, opportunities, expectations, expressions and identities of girls, women, boys, men, and gender diverse people. It influences how people perceive themselves and each other, how they act and interact, and the distribution of power and resources in society. Gender is usually conceptualized as a binary (girl/woman/femininity and boy/man/masculinity) yet there is considerable diversity in how individuals and groups understand, experience, and express it.” There are nuanced differences in how gender identity and presentation influences the development of CAD risk such as the onset of CAD risk factors, time to treatment and prognoses that are discussed in depth elsewhere (38) and not the focus of this review. Hereafter, mentions of “women” are assumed cisgender females with 46,XX chromosomes at birth, and “men” assumed cisgender males with 46,XY.

## Methods

### Search strategy

A scoping review of the literature was conducted in consultation with a librarian by doing parallel electronic searches of MEDLINE (via Ovid, 1946 – April 24, 2024) and EMBASE (via Ovid, 1974 – April 24, 2024). The initial part of the search strategy included synonymous terms related to our desired atherosclerotic coronary artery disease phenotype: “Coronary Artery Disease” or “coronary heart disease or coronary artery disease.ti”. The second part of the search strategy included terms related to our desired study type: “Genome wide association study.mp. or Genome-Wide Association Study/”. The phenotype and study type were linked using the “AND” operator. In addition, manual searching was conducted through citations of relevant reviews and cross-referencing with studies published in the GWAS catalog (https://www.ebi.ac.uk/gwas/home).

### Study selection

To determine eligibility for full-text review, title and abstract screening was conducted by two independent reviewers (A.A., E.T.). All conflicts were resolved through discussion with investigators with the relevant domain expertise (A.L., T.S.). The same applied for screening full-text articles. Studies included were those of: 1) CAD phenotype; and 2) original GWAS or a GWAS meta-analysis.

There is heterogeneity in how CAD is defined in the GWAS literature. In general, obstructive CAD refers to ≥50% stenosis in the left main coronary artery or ≥70% in a major epicardial vessel identified through coronary angiography (39). However, in studies where researchers do not include angiography results, a history of MI and/or percutaneous coronary intervention (PCI) and/or coronary artery bypass grafting (CABG) are often used as proxies for CAD. Therefore, we included papers with a definition of CAD as: 1) luminal stenosis more than 50% in a major coronary artery; 2) MI; 3) CABG and/or 4) PCI. An “original” GWAS refers to genome-wide association analyses conducted in a novel study population. This includes papers from consortia that published consecutive GWASs over time from cohorts that expanded in case sample size over time.

We excluded abstracts where no full text exists, non-English studies, non-GWAS study design (e.g. family studies, linkage analyses, candidate gene studies), CAD risk factors assessed as outcome (e.g. blood pressure, lipid traits), any non-CAD outcomes (e.g. heart failure), and any studies that exclusively were a secondary analysis using previously published GWAS data (e.g., Mendelian randomization, polygenic risk scores, causal pathway analysis).

### Data extraction

Following abstract screening, full texts were examined for further exclusion. We limited our analysis to any candidate gene with a corresponding single nucleotide polymorphism (SNP) rsID that had a P-value below the widely accepted Bonferroni-corrected genome-wide significance value of p < 5*10^-8^. Data corresponding to these SNPs were extracted from all materials available, including any supplemental materials.

For the studies that met the above criteria, full texts were reviewed for extraction of the following variables: Study identifiers (such as author, year of publication, title), study characteristics (sample size, CAD definition, genetic technology used to identify SNPs, population ethnicity(ies), sex ratio of included participants) and candidate gene information (SNP’s rsID, mapped gene(s), chromosomal position, risk allele, odds ratio, P-value, minor/estimated allele frequency). When sex/gender was regressed out as a covariate and not treated as an independent variable, these analyses were termed “sex-combined” analysis for our purposes. When quantifying ethnicity and ancestry representation, the following were grouped together: “White” and “Caucasian” as “European”; “Han Chinese”, “Japanese”, “Taiwanese” and “Korean” as “East Asian”; “Pakistani” and “Bangladeshi” as “South Asian”; “Saudi Arabian” and “Lebanese” as “Middle Eastern”; and “Black”, “Black British” and “African American” as “African descent”.

### Curating gene list from SNPs

A summary of all genes mapped from SNPs was made using information provided by study authors of which SNPs were mapped to genes. If a SNP was not previously mapped to (a) gene(s) by study authors, they were excluded from further consideration; we did not infer mapping so as to not introduce bias in the interpretation and inflate (e.g. as SNPs can be mapped to multiple genes). Standardization of gene names was conducted, and the number of times each gene was listed by authors were quantified. If a SNP was mapped to multiple loci as explicitly described in the publication, all genes were included. The purpose of this review was not to interpret putatively causal genes from mapped SNP loci towards CAD risk. However, recognizing how many SNPs may be mapped to inconsequential loci (from functional points of view), for studies that conducted non-sex-stratified analyses, a cutoff of genes appearing in at least 5 studies was considered for literature review of evidence of the gene’s relevant associations with coronary artery disease, and if there were any studies suggesting modulation by estrogens. Considering how few studies produced genome-wide significant sex-specific results, this minimum of 5 studies was not applied to the sex-stratified candidates.

## Discussion

### GWAS Study Characteristics

There were 36 studies that met inclusion criteria for review of gene candidates with genome-wide significant SNPs reported, including one paper that was included from manual searching (Figure 1). Key cohort characteristics of these studies are summarized in Table 1. Studies that met inclusion criteria but did not contain genome-wide significant results are summarized in Supplemental Table 1.

**Figure 1:**
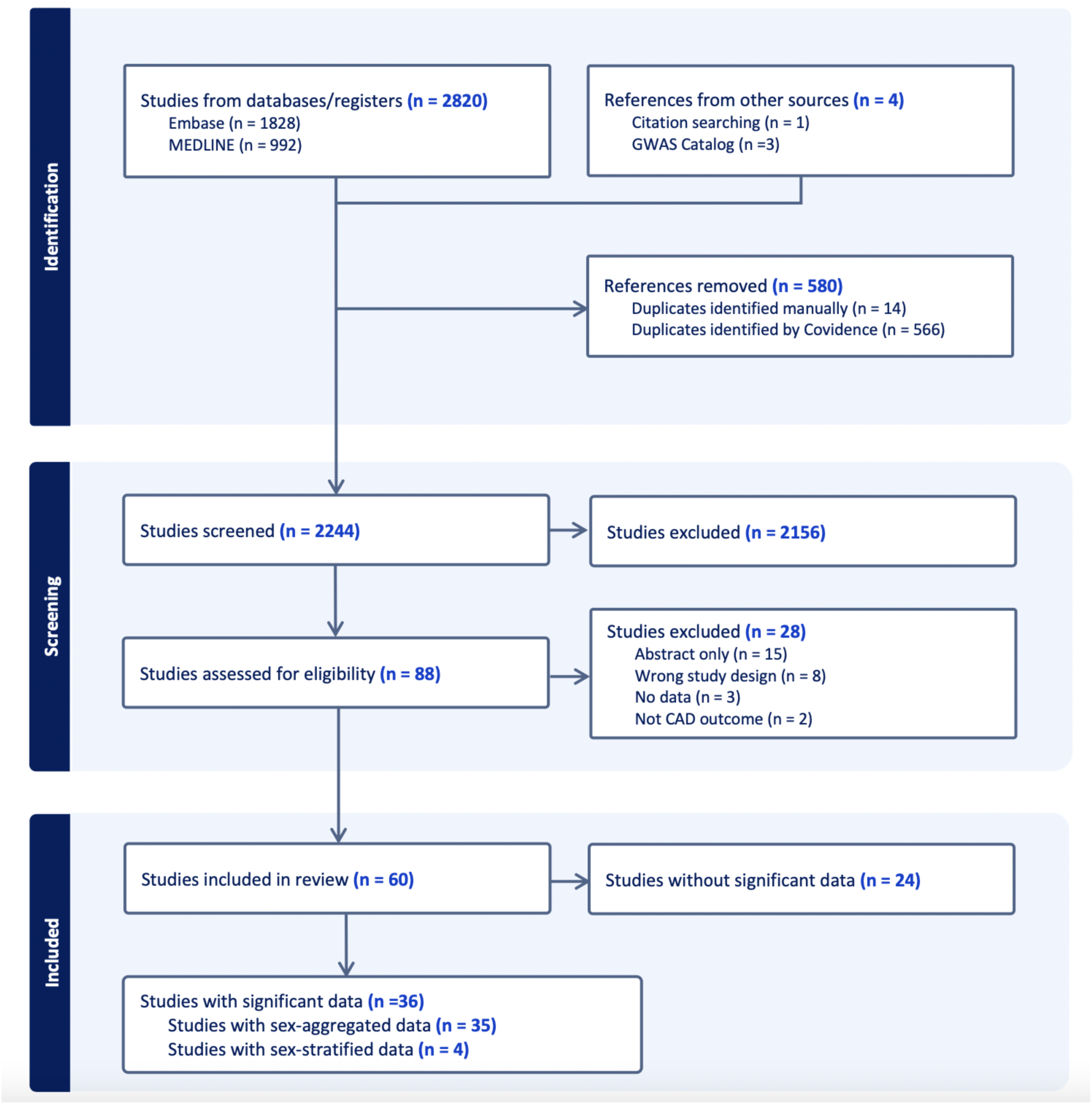
Abstract screening of CAD GWAS studies included shown through PRISMA flow diagram. “Significant” refers to genome-wide significance of p<5*10^-08^ and “data” refer to SNPs.

**Table 1:** Summary of GWAS studies included that had genome-wide significant data (n=36)

Of the 36 studies included, only 4 reported any genome-wide significant sex-stratified results, from whom 3 had mapped their results to genes (Figure 2A) (40–42). More details of the studies and SNPs identified from sex-combined and sex-stratified analyses can be seen in Supplemental Tables 2 and 3, respectively. The size and clinical heterogeneity of the CAD presentation within and between cohorts are suspected to contribute to the under-representation and lack of sex-specific results. However, exploring the reasoning for why women were under-enrolled in the cohorts used for these GWAS analyses is beyond the scope of this paper,

**Figure 2:**
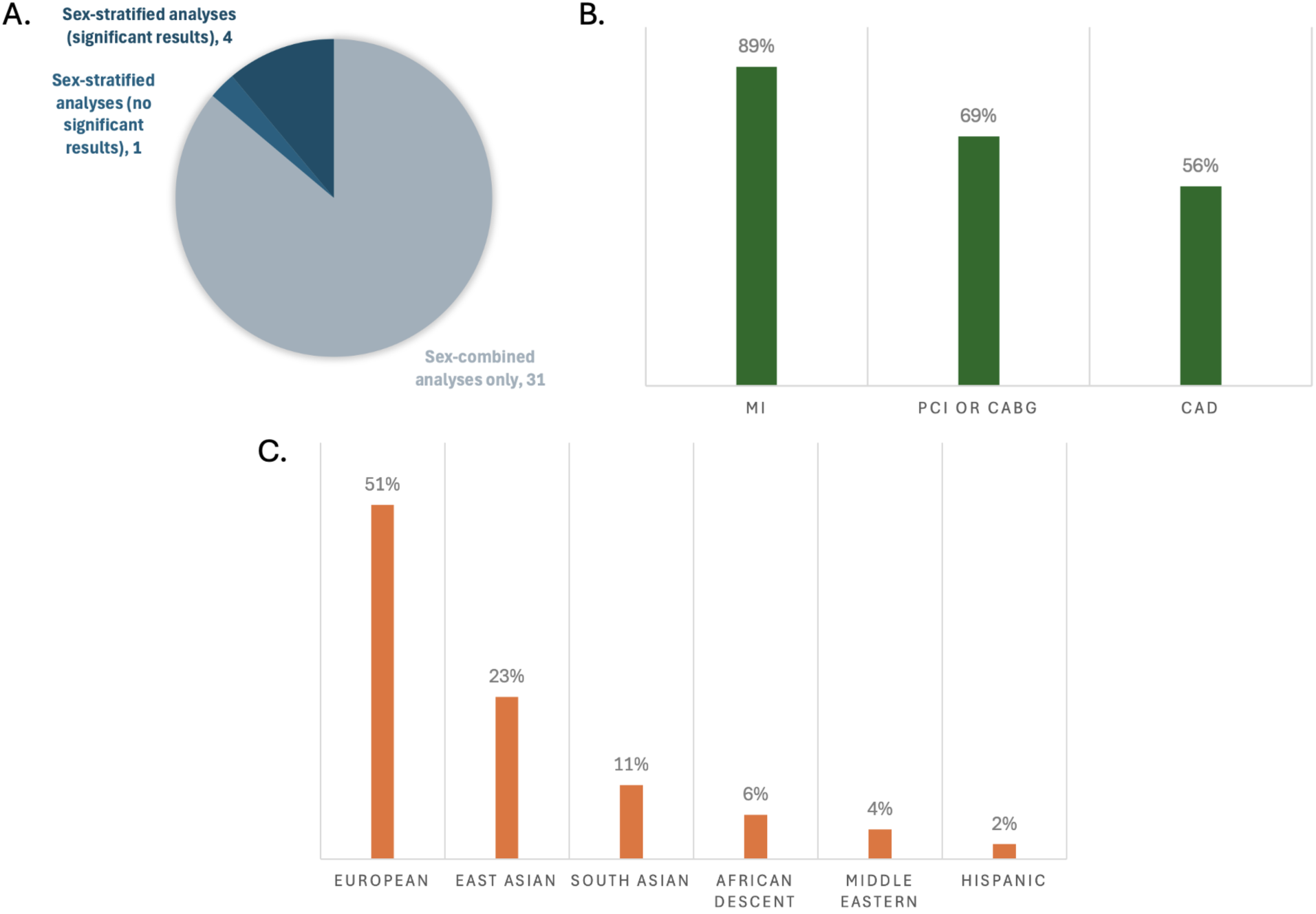
Representation of key characteristics of the GWASs that had genome-wide significant SNPs mapped to genes (n=36). A. Frequency of studies that did sex-stratified analyses and/or sex-combined analyses; B. Frequency of definitions used for cases; C. Frequency of inclusion of the large ethnicity supergroups in the studies. Note: Percents do not add up to 100% because multiple CAD definitions or ethnicities may have been reported per study.

**Table 2:** All SNPs with a p<5×10^-8^ mapped to genes that were replicated in at least 5 GWASs that did sex-combined analyses are presented here. A total of 324 unique significant SNPs were mapped to the 52 genes. 26 of these genes were found to have modulation or relation to estrogen pathways. 9 of these genes had evidence of estrogen modulation unrelated to CAD.

Most of the 36 studies included composite definitions that specified at least one of MI, PCI, CABG, and/or CAD with stenosis limits of >50%, >70% or >75%. In their definition of CAD, 33 studies included MI, 25 included CABG, 24 included PCI, and 20 included >50%, >70% or >75% stenosis limits (Figure 2B). An important limitation to underscore in any generalization of MI as CAD, in the absence of PCI, CABG or angiographic reporting of >50% epicardial stenosis, is the potential for confounding with non-atherosclerotic causes of MI. These causes are impossible to quantify without further individual-level information regarding the event. Specifically, MI with no obstructive coronary arteries (MINOCA) is a term encompassing coronary plaque disruption due to rupture, erosion, and calcific nodules (atherosclerotic causes) and non-atherosclerotic etiologies of MI, the most prevalent being epicardial vasospasm, coronary microvascular dysfunction (CMD), and spontaneous coronary artery dissection (SCAD). Further, non-ischemic causes of MI include MINOCA “mimickers”, such as Takotsubo cardiomyopathy, myocarditis and supply-demand mismatch (Type 2 MI) (43). MINOCA makes up about 6-15% of all MIs, (43),and is approximately 3 times more prevalent in women than men (44), which adds to clinical heterogeneity in CAD groups solely using MI as their inclusion criteria. Interestingly, a high polygenic risk score (PRS) for SCAD has been shown to have associations with lower risk for atherosclerotic CAD and vice versa (45). Therefore, robust phenotype inclusion for CAD GWAS study designs are recommended so as to not risk accidental inclusion of individuals with non-CAD etiologies of MI, which could influence statistical significance of SNP candidates.

Most studies (26, 72%) reported only one ethnicity. The most commonly reported individuals were of self-reported European ethnicity (51%), followed by East Asian (23%), then South Asian (11%) (Figure 2C). There is a plethora of research demonstrating epidemiological differences in CAD prevalence in different racial/ethnic subgroups, for example with 2-4x higher rates observed in South Asian (46,47), Hispanic, and Black (48) individuals compared to White individuals. There is an ongoing challenge by researchers to disentangle genetic risk (ancestry) from social (race/ethnicity) and environmental influences on this risk, such as differences in diet, exposure to stress, exercise, that increase CAD risk through variation in gene expression that strongly affect these observed ethnic differences (49) (Box 2). The generalizability of results produced by SNP array technologies used in GWASs should be scrutinized based on the population for which the arrays were designed; for example, accuracy is greatly reduced when SNP arrays created from European-ancestry allele frequencies are applied on individuals of African ancestry (50). This technical bias is further perpetuated by analyses that include filtering of alleles of rare frequency (typically <5% or <1% prevalence in a referenced population). Importantly, there is the concept of “sex-influenced inheritance” (a.k.a. “sex-biased inheritance”) wherein there are more inter-sex similarities than inter-population similarities for a number of traits, reflected in sex-stratified analyses comparing the sexes within cohorts of diverse genetic ancestry. This is exemplified in recent very large multi-ethnic sex-stratified GWASs of complex traits such as blood pressure (51) and lipid traits (52,53) identifying sex-specific gene associations contributing to the traits. The more that risk genes are identified and replicated between GWAS studies with sex-stratified analyses of diverse genetic ancestries, the more robust these genes are for generalizability across population groups.

#### Box 2

In health research, the concepts of “race”, “ethnicity” and “genetic ancestry” are terms often used interchangeably when referring to inter-population differences in prevalence and outcomes of disease observed between groups. In genomics research, differences in SNP prevalence between these groups has implications when evaluating rarity against a background referent population in databases such as the Genome Aggregation Database (https://gnomad.broadinstitute.org/). However, race/ethnicity are social constructs and do not reflect biological variation underpinning these differences as much as the term “genetic ancestry” attempts to do so. Race is a construct used to categorize people based on perceived differences in physical appearance, such as skin tone (54) whereas ethnicity refers more to the community and shared cultural group membership with features such as shared language, geographic origin, nationality, cultural traditions, migration history, etc. (54). Genetic ancestry refers to the inheritance of segments of DNA from “source” populations; the majority of individuals contain a mosaic of ancestries from different populations (“admixed”), and this heterogeneity does not always correlate with self-described or perceived race/ethnicity (50).

### Gene Candidates and Estrogen Modulation

From the 36 studies included, 61 genes were identified for further literature review if modulated by estrogen. There were 52 genes found in sex-combined analyses with corresponding SNPs surpassing genome-wide significance in at least 5 studies (Table 2), and 29 genes found in sex-stratified analyses whose corresponding SNPs also surpassed genome-wide significance (Table 3). Due to the small fraction of studies that had significant SNPs mapped to genes from sex-stratified analysis (n=3), all genes that surpassed genome-wide significance were retained, even if not replicated. The majority of these 29 sex-stratified gene associations (n=20/29) were also identified in the sex-combined analyses replicated in at least 5 other studies (Figure 3).

**Figure 3:**
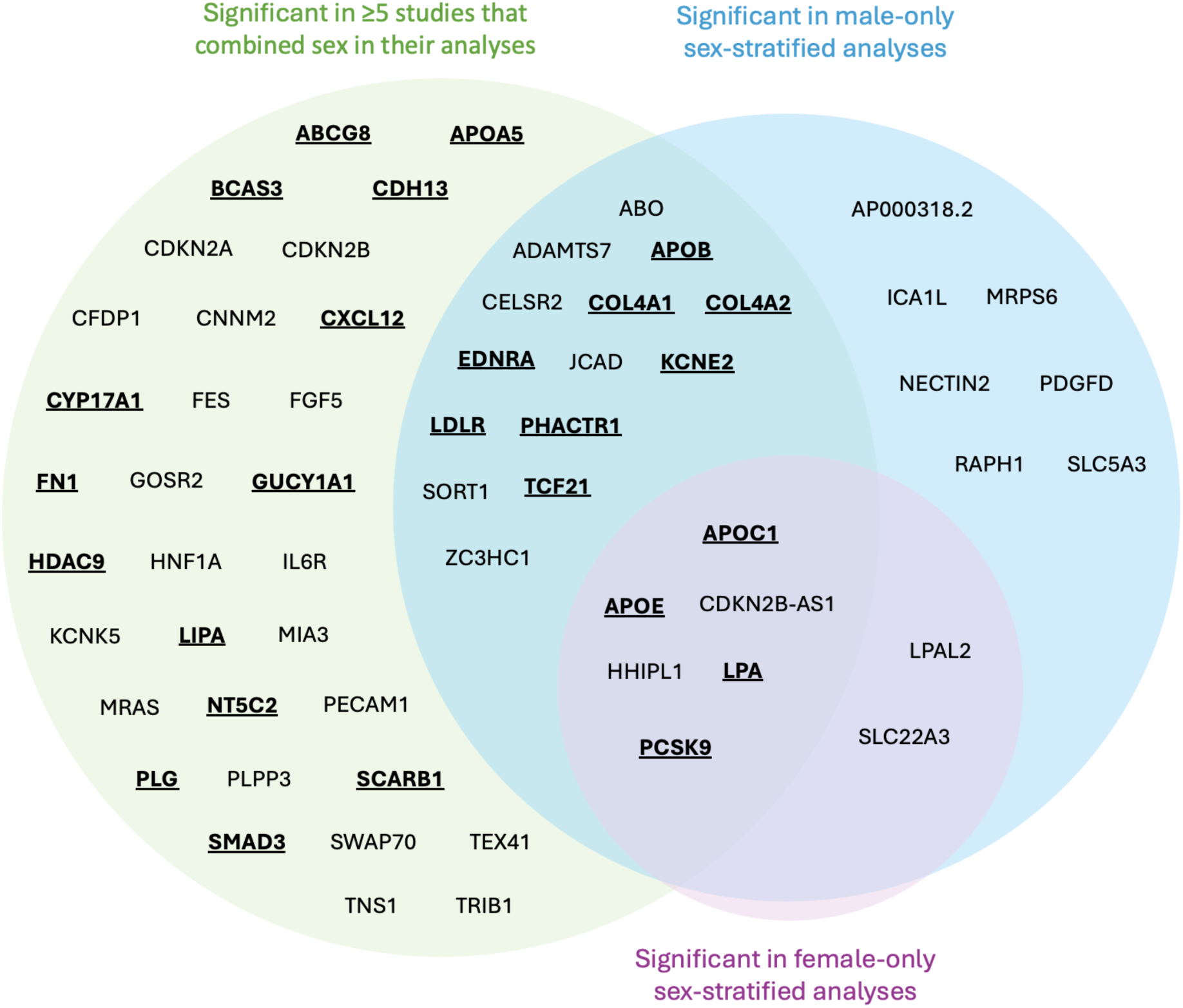
Venn diagram mapped genes from significant SNPs identified in CAD GWASs. Genes emphasized by bold and underscore were identified to have evidence of estrogen modulation of gene expression and/or translated protein activity. No SNPs were identified to be significant just in females.

**Table 3:** All SNPs with a p<5×10-8 mapped to genes found in at least 1 sex-stratified GWAS are presented here. A total of 26 unique significant SNPs were found among the 29 genes. Of these 29 genes, 20 of them were also identified in sex-combined analyses. 21 genes were identified in male sex-stratified analyses. No genes were found to be significant in females only that weren’t also identified in male sex-stratified analyses. 12 were found to have modulation or relation to estrogen pathways. 4 of these genes had evidence of estrogen modulation unrelated to CAD.

Following literature review of the 61 genes, 26 (43%) were identified to be affected by estrogen (Figure 3, bold and underlined text). All 26 genes were identified in at least 5 papers with sex-combined analyses, while 12 of these 26 were also identified in at least 1 sex-stratified analysis. Of these 12, 8 were identified in male-stratified analyses in addition to the sex-combined analyses (*APOB, COL4A1, COL4A2, EDNRA, KCNE2, LDLR, PHACTR1, TCF21*), and 4 were identified in the male-stratified, female-stratified, and sex-combined analyses (*APOC1, APOE, LPA, PCSK9*). Of note, all significant sex-stratified gene associations identified in females also overlapped with males. None of these 26 genes are on the X-chromosome. The remaining 35 genes did not have literature supporting any modulation of estrogen, directly or indirectly through relation to CAD or other diseases (e.g., breast cancer). These 35 genes were: *ABO, ADAMTS7, AP000318.2, CDKN2A, CDKN2B, CDKN2B-AS1, CELSR2, CFDP1, CNNM2, FES, FGF5, GOSR2, HHIPL1, HNF1A, ICA1L, IL6R, JCAD, KCNE5, LPAL2, MIA3, MRAS, MRPS6, NECTIN2, PDGFD, PECAM1, PLPP3, RAPH1, SLC22A3, SLC5A3, SORT1, SWAP70, TEX41, TNS1, TRIB1,* and *ZC3HC1*.

Of the 26 genes with evidence of estrogen modulation, the connection with CAD can be largely categorized by their involvement in lipid production/metabolism, arterial vascular properties, and/or thrombosis, and are expanded on below with relation to genetic variation towards CAD risk.

### Lipids

The following 12 genes were classified as having a role in lipid synthesis, metabolism and/or lipoprotein mechanisms: *LDLR, APOB, PCSK9, LPA, LIPA, ABCG8, SCARB1, APOA5, APOE, APOC1, KCNE2,* and *HDAC9.* Of these, most had evidence of estrogen modulating some aspect of the CAD-related process, except *KCNE2* and *HDAC9*, which had estrogen modulation evidence in other tissues (e.g. breast tissue, ovarian tissue).

*LDLR* encodes a cell surface receptor predominantly expressed in the liver involved in the endocytosis of LDL particles, where they are further metabolized and degraded in hepatocytes (55,56). Missense or loss of function (LoF) mutations in the *LDLR* gene results in reduced affinity or inability of the LDLR to bind LDL, resulting in higher plasma lipid levels and thus increased risk of hypercholesterolemia (57). Estrogen influences plasma lipid concentration through LDLR-dependent and LDLR-independent pathways (58). It has been shown in rabbit and rat models that LDLR expression increases up to 10-fold when treated with estrogen, resulting in increased clearance of plasma LDL (59,60).

*APOB* encodes for a major apolipoprotein ApoB, which is a high affinity ligand for LDLR attached to LDL particles (61). Missense or LoF variants in *APOB* can result in decreased binding affinity of LDL to bind to LDLR, resulting in elevated plasma lipid levels (62). Estrogen can reduce ApoB concentrations as demonstrated by experiments in mice via LDLR-independent mechanisms (63).

*PCSK9* encodes proprotein convertase subtilisin/kexin type 9, which plays a crucial role in cholesterol metabolism by promoting LDLR degradation (64). *PCSK9* gain of function (GoF) mutations further increase LDLR, thus increasing plasma lipid levels. Estradiol reduces PCSK9-mediated LDLR degradation through a mechanism involving activation of the G-protein-coupled estrogen receptor (65). Moreover, circulating levels of PCSK9 are generally higher in women than in men, and this difference is more pronounced post-menopause, which has been suggested to be due to lower circulating estrogen(66). However, in premenopausal women, PCSK9 levels vary with the menstrual cycle, showing an inverse relationship with estradiol (67).

Several studies of the *SLC22A3-LPAL2-LPA* gene cluster have suggested that polymorphisms in this region are associated with an increased risk of CAD (68,69). From our review, among genes with evidence to be modulated by estrogen, the SNPs mapped to genes in this cluster had the highest odds ratios, with the highest in *LPA* of 1.51 (1.33-1.70) (Refer to Supplementary Table 2).

*LPA* encodes a highly polymorphic glycoprotein, apolipoprotein-a (“apo-a”), which attaches to LDL and is, together, referred to as lipoprotein-a (“Lp(a)”). Lp(a) is similar to LDL but an independent risk factor of CAD that does not lower with statins (70). Copy number variations in the kringle-iv repeat region of *LPA* are the main genetic determinants of circulating Lp(a) concentrations (71). When apo(a) is cleaved, it results in fragments that are prone to attaching to atherosclerotic lesions and promoting thrombogenesis through plasminogen activation (70,71). There is evidence that estrogen decreases Lp(a) plasma levels by increasing its uptake via LDLR (72). In postmenopausal women, taking hormonal therapy has been associated with lower Lp(a) values and subsequent lower risk of CAD (72).

*ABCG8* encodes for jejunal and ileal sterol efflux transporters (73). LoF mutations of *ABCG8* have been shown to lead to elevated plasma sitosterol and LDL levels (74). ABCG8 has been shown to significantly increase sitosterolemia and potentially accelerate progression of CAD (75). Estrogen upregulates intestinal *ABCG8* activity through the intestinal ERa pathway, thus leading to increased cholesterol absorption (73). Interestingly, this upregulation can be completely counteracted by use of an estrogen antagonist (73).

*APOA5* encodes a minor apolipoprotein that is an important component of the high density lipoprotein (HDL) and very lower density lipoprotein (VLDL) (76). It directly regulates triglycerides by increasing lipoprotein lipase (LPL), which stimulates the breakdown of triglyceride-rich lipoproteins resulting in lower plasma triglycerides levels (77). Knockout mouse models of *APOA5* have shown 4-fold increases in triglyceride levels, and as such it has been linked as a CAD risk gene (78). Oral administration of estrogen has been shown to increase triglyceride concentration, suggested to be due to increased hepatic production of triglycerides due to reduced ApoB production and decreased triglyceride clearance through inhibition of LPL (79). In addition, APOA5 has been shown to be higher in women than in men, potentially suggesting that there may be sex differences in its regulation (80).

*APOE* encodes a plasma protein that is involved in transport and metabolism of cholesterol and triglycerides (81). It is a major ligand for LDLR (82) and is known for having 3 major alleles impacting disease risk: APOE2, APOE3, and APOE4 (82). Expression of these alleles result in higher circulating plasma LDL levels: the E2 and E3 alleles demonstrate poorer binding affinity to LDLR receptors, and E4 alleles bind preferentially to very low density lipoprotein particles (VLDLs), resulting in decreased lipid uptake by LDLR on hepatocytes (82,83). While these variants have similar background population frequencies between sexes, there have been sex differences observed in associated cardiovascular risk. For example, the E4 allele is associated with higher risk in men, while the E2 has a protective effect observed in women only (84). *APOE* has been demonstrated to be upregulated by estrogen through the estrogen-receptor alpha pathway (85), although the effects are variable based on the *APOE* alleles carried. For example, the E4 allele has shown to have higher expression following menopausal hormone therapy than E2 and E3 variants (84). Many studies use APOE-knockout murine models to study accelerated atherosclerosis and effects of estrogen, underscoring the importance of this ligand in atherosclerotic pathology (86).

*SCARB1* encodes an HDL receptor that mediates the cholesterol transfer to and from HDL (87). LoF or missense alleles increased dimerization and decreased hepatocellular uptake of HDL, resulting in higher risk of atherosclerosis (87). Estradiol has been shown to be an indirect modulator of the SCARB1 receptor in rat models (88). Estradiol does not directly decrease expression of *SCARB1* in the liver, however it has been demonstrated to decrease *SCARB1* expression levels secondary to the estrogen-induced increase in LDLR activity and ACTH presence (88).

*LIPA* encodes lipase A, which functions in lysosomes to hydrolyze cholesteryl esters and triglycerides to generate free cholesterol and free fatty acids following LDLR-mediated LDL endocytosis in hepatocytes (89). LoF variants in LIPA have been shown to result in accumulation of triglycerides and cholesterol esters that contribute to foam cell development and premature atherosclerotic plaque formation (89,90). Deficient *LIPA* activity may have an indirect effect on CAD risk through its downstream effects on estrogen production. Since LIPA functions in hydrolysis of triglycerides and cholesterol, it provides energy while also influencing the synthesis and secretion of sex hormones including estrogen (91).

*APOC1* encodes an apolipoprotein C1 family member that plays a role in HDL and VLDL metabolism through highly selective inhibition of cholesteryl ester transfer protein (CETP) in plasma (92,93). Decreased expression of APOC1 from LoF or missense variants, leads to increased plasma triglycerides, which confers increased CAD risk (94). One study suggested that APOC1 could promote the estrogen receptor expression in the context of ovarian cancer (95). In addition, there is marked APOC1 elevation in women with PCOS; however it is unclear how much of the altered lipid metabolism is due to estrogen and/or androgen metabolism or because of insulin resistance (96).

*KCNE2* encodes a voltage-gated potassium channel involved in regulating heart rate, neurotransmitter release and smooth muscle contraction (97). Demonstrated by mouse knockout models, causal links between *KCNE2* and atherosclerosis have been suggested to be due to raising serum LDL and impairing glucose tolerance (98,99). Estrogen action alters *KCNE2* expression directly through binding of estrogen receptor alpha to the estrogen-responsive element in the *KCNE2* regulatory domain (100). Estrogen was observed to be less responsive in the presence of variants altering affinity of DNA-binding domains in either the estrogen receptor or *KCNE2* estrogen-responsive element (100). Further, the nuclear estrogen-related receptor has also been shown to have a crucial role in modulating *KCNE2* by binding to its promoter (101).

*HDAC9* encodes enzyme histone deacetylase 9, and has been implicated in progression of atherosclerosis through histone acetylation and subsequent expression of specific genes related to lipid metabolism and macrophage polarization (102). Missense and LoF *HDAC9* mutations result in downregulation of inflammatory genes and increased Apolipoprotein-A1 and HDL-mediated cholesterol efflux, resulting in decreased plasma cholesterol levels (103). As such, it is thought that upregulation of *HDAC9* in macrophages has increased atherosclerotic risk through suppression of cholesterol efflux and proinflammatory actions (103). A variant has also been shown to directly modulate the expression of *TWIST1*, a gene that regulates arterial wall proliferation and calcification (104). Increased expression of *HDAC9* has been shown to be associated with decreased expression and activity of estrogen receptor alpha in MCF-7 cells in breast cancer studies (105).

### Vascular Integrity

The following 11 genes were classified as having a role in vascular integrity through roles in the vascular endothelium and/or smooth muscle cells: *BCAS3, COL4A1, COL4A2, SMAD3, CYP17A1, CDH13, CXCL12, EDNRA, NT5C2, TCF21, PHACTR1*. Of these, *BCAS3, COL4A1, COL4A2, SMAD3* and *CYP17A1* had evidence of estrogen modulating some aspect of CAD development, and the rest had estrogen modulation evidence in other tissues (e.g. breast tissue, uterine endothelium).

*BCAS3* encodes for a cytoskeletal protein that functions in angiogenesis and related processes like TGFβ signaling, cell adhesion, peptidase activity and matrix organization (106). It is thought to do this by activating Cdc42 which in turn affects actin organization, cell polarity and cell motility in endothelial cells (107). Endothelial *BCAS3* knockout mice survive to only embryonic day 11.5 and have diffuse vascular patterning defects (106). However, despite numerous studies citing *BCAS3* association with CAD, functional studies are still lacking. Estrogen directly induces *BCAS3* transcript expression by binding to estrogen receptor alpha and subsequently inducing *MTA1*, which is a transcriptional factor of *BCAS3* (108).

*COL4A1* and *COL4A2* encode subunits of type IV collagen, which is an essential structural component of the basement membrane (109). LoF alleles in these genes result in lower collagen IV abundance and thinner fibrous caps, thus creating very unstable plaques and contributing to smooth muscle cell pathology (110). *SMAD3* encodes the SMAD family member 3, a signaling molecule involved in the transforming growth factor-beta (TGF-β) signaling pathway(111), which is a cell growth inhibitor crucial in the regulation of inflammation and fibrosis (112). *SMAD3*-knockout mice have decreased fibrotic response, resulting in thin, unstable fibrous caps (113). SMAD3 induces TGF-β which is necessary for TGFβ-stimulated expression of both *COL4A1* and *COL4A2* (113). Estrogen inhibits TGF-β signaling by binding to estrogen receptor alpha, thus decreasing SMAD3 levels, which subsequently decreases *COL4A1* and *COL4A2* expression (114,115).

*CYP17A1* encodes a member of the CYP450 superfamily of enzymes, which contain a heme cofactor and mostly function as monooxygenases (116). CYP17A1 is specifically localized in the endoplasmic reticulum and involved in the steroidogenic pathway that produces mineralocorticoids, glucocorticoids, androgens, progestins and estrogens (117). One study showed that *CYP17A1* knockout mice develop atherosclerotic lesions at a higher rate compared to WT (118). The functional evidence linking *CYP17A1* and CAD is not entirely understood, however it is thought to involve glucose homeostasis regulation by promoting glucose uptake and utilization (118). Further, there is evidence of genetic variants that are associated with severe hypertension, a well-known risk factor for CAD (119,120). CYP17A1 has a role in estrogen production; it is highly expressed in granulosa cells, and can catalyze pregnenolone and progesterone to form androstenedione, which CYP19A1 later converts to estrogen (121). Missense/LoF variants in *CYP17A1* have been shown to cause fertility impairments and related cancers (119,122).

*CDH13* encodes a T-cadherin that is expressed on endothelial and smooth muscle cells (123). Increased CDH13 activity results in increased migration and proliferation of endothelial cells and therforefore effects vascular remodeling and atherosclerosis development (124). Knockout T-cadherin and adiponectin mice had increased neointimal thickness following carotid artery ligation (125). A variant within this gene was found to be associated with estrogen signaling metabolism in breast cancer, menstruation patterns and pregnancy (126).

*CXCL12* encodes for a chemokine that is produced in endothelial cells and as such has an important role in angiogenesis, hematopoiesis, and tissue regeneration (127,128). *CXCL12*-knockout mice models demonstrated compromised artery coverage, suggesting a key role in arterial development and regulation (129). CXCL12 has also been shown to consistently be upregulated in individuals with calcific aortic valve disease (130). However, the underlying mechanism between poor prognosis of CAD and high levels of CXCL12 is not understood (127). Research has shown estradiol regulation of the CXCL12 axis in the growth of breast cancer cells (131). Estradiol directly induces transcription of *CXCR4* and *CXCR7*, which are both receptors of CXCL12, through control of their promoters (131).

*EDNRA* encodes endothelin receptor A, which mediates cell proliferation and long-lasting vasoconstriction (132). In coordination with EDNRB, EDNRA can mediate contraction post-relaxation of intact endothelium vessels (132). Increased *EDNRA* expression has shown to play an important role in hypertension and thus progression of vascular proliferation (132,133). One study showed that *EDNRA* transcript in endometrial stromal cells increases in the proliferative phase when given estradiol in rhesus macaque (134). Estrogen binding to estrogen receptor alpha directly results in increased epithelial *EDN3* expression, which is thought to act via EDNRA to further stimulate cell proliferation (134).

*NT5C2* encodes for a 5’ nucleotidase that functions in purine metabolism (135), and has a suggested role in type 2 diabetes and hypertension (136). NT5C2 is present on vascular endothelium cells and likely has an inflammatory process relation to CAD (136,137). Knockdown *NT5C2* zebrafish had higher blood flow and elevated linear velocity, in addition to increased inflammatory markers such as angiotensin-converting enzyme and C-reactive protein (138). There is some evidence to suggest that NT5C2 may be involved in the estradiol regulation of fibroblasts (137). Expression levels of *NT5C2* were increased 2 hours after estradiol administration in primary uterine endometrial epithelial cells (137).

*TCF21* encodes a transcription factor that has been identified as a “master regulator” for smooth muscle cell gene expression (139). Increased *TCF21* expression has been suggested to be associated with decreased risk of CAD by influencing smooth muscle cell behavior in developing lesions, contributing to a protective fibrous cap (140). It achieves this by disrupting the MYOCD-SRF pathway, which is crucial for SMC differentiation (140), as shown by in vitro experiments of variants causing overexpression of *TCF21* and *TCF21*-knockout mouse models (141). TCF21 interacts with USF2 in endometriotic stromal cells and activates the promoters of SF-1 and estrogen receptor beta, thereby influencing the estrogen pathway in endometriosis While it is unknown if this estrogen activation is widespread throughout the body, it is interesting to note that TCF21 also regulates fibrosis in endometriosis (142).

*PHACTR1* encodes for a member of the phosphatase and actin regulator family It has a crucial role in binding to actin to regulate the organization of the actin cytoskeleton and important roles in tubule formation, and thus in endothelial cell survival (143). *PHACTR1* deficiency from knockout mice or LoF experiments demonstrated accelerated foam cell formation and thus increased atherosclerosis (144). In ovarian granulosa cells, *PHACTR1* was identified to be a regulatory target by estrogen through experiments in estrogen receptor 2-depleted mice (145); however this connection has not been replicated in vascular endothelial cells.

### Thrombosis

Three genes were identified to have a role in thrombosis, and all had evidence of estrogen modulating some aspect of CAD development: *GUCY1A1, PLG* and *FN1*.

*GUCY1A1* encodes the alpha subunit of the guanylate cyclase enzyme, which is an essential enzyme in platelets (146). LoF alleles result in lack of guanylate cyclase enzyme in platelets, which have been shown experimentally to result in vascular inflammation through increased leukocyte recruitment and endothelial cell activation, and thus atherosclerotic plaque progression (146). Estradiol has been shown to highly up-regulate both expression and activity of the a1 subunit of guanylate cyclase through estrogen receptor activation (147).

*PLG* encodes a plasminogen protein that is converted to active plasmin by plasminogen activators such as tissue plasminogen activator (148). Plasmin acts as an antithrombotic agent, and is responsible for degrading fibrin-containing blood clots (148), in addition to cleaving fibronectin and von Willebrand factor (149). It is hypothesized that increased binding of plasminogen kringle domains to tissue plasminogen activator can lead to increased activator activity, thus increasing the conversion of plasminogen to plasmin, and as such cause unstable plaque formations to occur (148). Estrogen directly increases *PLG* expression by binding to 5’-region-flanking enhancers, and is tissue-specific to hepatocytes (150).

*FN1* encodes fibronectin, a glycoprotein involved in the cell adhesion and migration processes of thrombosis and coagulation (151). Fibronectin works with fibrin and fibrinogen in clot formation, contributing to thrombus stability (151). Fibronectin has also shown to play a role in mediating platelet adhesion(151). In knockout mouse models, a splice variant of fibronectin containing extra domain A has been shown to have twice the amount of atherosclerotic lesions and macrophage content in plaques (152). Inhibition of the estrogen receptor is suggested to significantly decrease *FN1* expression (153), consistent with studies demonstrating increased expression in breast cancer cells through G-protein coupled transmembrane receptors (154).

### Limitations

There are several limitations from the methods used in our study. It must be acknowledged that due to the lack of sex-stratified data available, sex-stratified gene associations were only required to be discovered once. This means that there is less credibility to these sex-stratified genes. This is in comparison to sex-combined genes analysis where a cutoff of 5 studies in which a gene was replicated was included. This cutoff was chosen so as to increase confidence in the SNPs included in the discussion, as several identified papers listed all potential genes that could be mapped to a SNP (41,155–157), without secondary analysis that would result in one high confidence causal gene.

Additionally, due to the nature of our search for estrogen, it is possible that there are connections between estrogen and genes that were missed. For example, any downstream metabolites or effectors of the estrogen system were not included in this review. In addition, estrogen is not the only sex hormone. There are androgens that may have a role in CAD that were not evaluated. Androgen regulation has demonstrated potential involvement in the CAD pathway (158), and is an area that continues to grow with more research.

Moreover, there are flaws innate to the study design of GWASs that limit the conclusions that can be made. To increase power in a GWAS, meta-analyses will incorporate many cohorts to increase sample size. However, this causes heterogeneity and leads to decreased confidence as many of these cohorts use varying inclusion/exclusion criteria, technology to identify SNPs, and have varying amounts of male to female participants. In fact, every study that reported case/control sex ratios had a larger percentage of men in the cases (data not shown) and more women being included as controls. We speculate that this may be due to the observation that women tend to develop CAD later in life than men by about 10 years (159) and/or assumptions that there are no genetic sex differences that will influence results. This may thus confound the validity of the results. It is thus an important consideration for future studies to include equal proportions of the sexes and conduct sex-stratified analyses to be able to identify candidates with sex-specific effects in the pathogenesis of CAD.

### Conclusion

Our scoping review identified 26 genes through CAD GWASs that are modulated by estrogen. Some of these genes are well-studied contributors to the development of CAD, such as *LDLR* and *PCSK9*. This study provides further evidence of the relationship between the actions of estrogen and the development of CAD. More research will need to be conducted to establish estrogen’s relation to sex differences in the pathology and progression of CAD.

## Data Availability

All data produced in the present work are contained in the manuscript and supplemental material.

## Author contributions

AA: Writing: original draft, review & editing. ET: Project conception, writing: original draft, review & editing. EB: Writing: review & editing. CKA: Writing: review & editing. AL: Writing: review & editing. TS: Writing: review & editing.

## Acknowledgments

We would like to thank UBC Faculty of Medicine librarian Aubrey Geyer for providing support during the design and execution of scoping review study methodology in the context of our article. The authors confirm that ethics protocol and patient consent forms were not required for this manuscript.

## Supplemental Tables

**Supplemental Table 1:** These 24 studies had sex-combined analyses that met all other inclusion criteria, but contained no SNPs that surpassed genome-wide statistical significance of p<5*10^-8^.

**Supplemental Table 2:** This table contains the 1658 SNPs that achieved genome-wide significance (p<5*10^-8^) from the included GWASs in sex-combined analyses.

**Supplemental Table 3:** This table contains the 597 SNPs from sex-stratified analyses that had achieved genome-wide significance (p<5*10^-8^) from the 4 papers that met our criteria. 413 SNPs were mapped from female-stratified data while 184 SNPS were mapped from male-stratified data.

